# Projecting international mpox spread in Asia: ongoing global health risk

**DOI:** 10.1101/2024.04.17.24305832

**Authors:** Toshiaki R. Asakura, Sung-mok Jung, Hiroaki Murayama, Cyrus Ghaznavi, Haruka Sakamoto, Ayaka Teshima, Fuminari Miura, Akira Endo

## Abstract

The global mpox outbreak affected many Asian countries in 2023, following a sustained local transmission in Japan. Given the large population sizes and limited vaccine rollout in Asia, the potential risk of global mpox reemergence arising from Asia is of concern. Using a mathematical model incorporating heterogeneous sexual networks among MSM, calibrated to incidence data in Japan, we projected the patterns of international mpox spread across 42 Asian countries. Our simulations highlight countries at a high risk of mpox introductions, many of which were low- and middle-income countries (LMICs) in South-eastern Asia. Our analysis also suggests a shifting focus of importation risk from Eastern Asia to South-eastern Asia, and subsequently to Central, Southern and Western Asia, which roughly coincided with the observed spread patterns in 2023. Global cooperation and support are warranted, especially for LMICs with an elevated risk of mpox introduction, to minimise the risk of continued circulation in Asia and beyond.

## Main text

While the global mpox outbreak rapidly spread among men who have sex with men (MSM) in many countries in early 2022, particularly in Europe and the Americas, Asia had been the least affected region with only limited numbers of importation and onward transmission reported by the end of 2022. However in 2023, following Japan’s local outbreak beginning in January, several countries in Eastern and South-eastern Asia experienced sustained transmission of mpox, notably China, which has reported the largest epidemic in the region with over 1,500 cases.^1^ Despite the World Health Organization (WHO) lifting the public health emergency of international concern (PHEIC) for mpox on 11 May 2023,^2^ there remain many Asian countries with large susceptible populations at risk, most of which are low- and middle-income countries with limited access to vaccines.

In the present study, we developed a mathematical model of mpox transmission over sexual contact networks among MSM to discuss and project the international spread patterns of the disease in Asia.

### Simulated importation probabilities in Asian countries

We constructed a stochastic meta-population SEIR model representing MSM populations in 42 Asian countries/territories. As the availability of MSM population size estimates substantially varies among these countries, we assumed that MSM account for one percent of the total population in every country considered.^3^ To account for the heterogeneity in sexual behaviours driving mpox transmission dynamics,^4^ the population in each country was stratified into 50 compartments of different sexual activity levels (reflecting the number of sexual partners over the assumed infectious period of mpox of 10 days^5^), which we assumed to linearly correlate with both the hazard of infection and onward transmission. We used the reported 4-week partner numbers from the British National Surveys of Sexual Attitudes and Lifestyles (Natsal) to parameterise the distribution of sexual activity levels among MSM populations in Asia, given the scarcity of similar datasets in this region. As an assessment of generalisability, we compared Natsal data against the National Inventory of Japanese Sexual Behavior (NInJaS) data^6^ in *Supplementary materials*. Mpox importations between countries were modelled based on the 2019 travel volume data from the United Nations World Tourism Organization (UNWTO).^7^ We assumed that the importation of mpox from one country to another could be contributed to by either inbound or outbound travellers, and that its risk was proportional to the number of travellers between the countries.

We simulated the international spread of mpox in Asia using a Bayesian data assimilation approach.^8^ We first fitted our model to the reported mpox incidence in Japan from 16 January (date of medical attendance for the first case in 2023) to 26 June 2023 using the approximated Bayesian computation^8^ to obtain posterior samples of the sexually-associated secondary attack risk (SAR; transmission risk per sexual partner) for mpox. For consistency with the observed local transmission in the Republic of Korea (South Korea) as of the end of the fitting period (26 June 2023), we also conditioned the posterior samples on the early introduction to South Korea; i.e. the model was required to have 10 or more cases in South Korea by 26 June 2023. We then simulated mpox transmission from 16 January onwards in Japan and other Asian countries with these posterior samples and the international travel volume data,^7^ assuming for simplicity that cases in Japan initiated the international mpox spread in Asia in 2023 and that importations from outside Asia were negligible. The posterior median estimate of SAR was 72.0% (95% credible interval: 47.2, 93.0), which was comparable to the observation from a contact tracing study in Belgium during the 2022 outbreak.^9^ Further methodological details can be found in *Supplementary materials*.

We computed the simulated importation probabilities for each country, which we defined as the proportion of simulations where the country experienced at least one importation of a case (Figure 1). Countries with the highest simulated importation probabilities mostly concentrated in Eastern and South-eastern Asia. Of note, 6 countries (mainland China, Thailand, Vietnam, Philippines, Malaysia, and Indonesia) among the top 10 were low- and middle-income countries (LMICs); likewise, 13 among the top 20 were LMICs. We also found that many of these countries indeed observed mpox introductions in 2023 (Figure S8).

**Figure 1.**
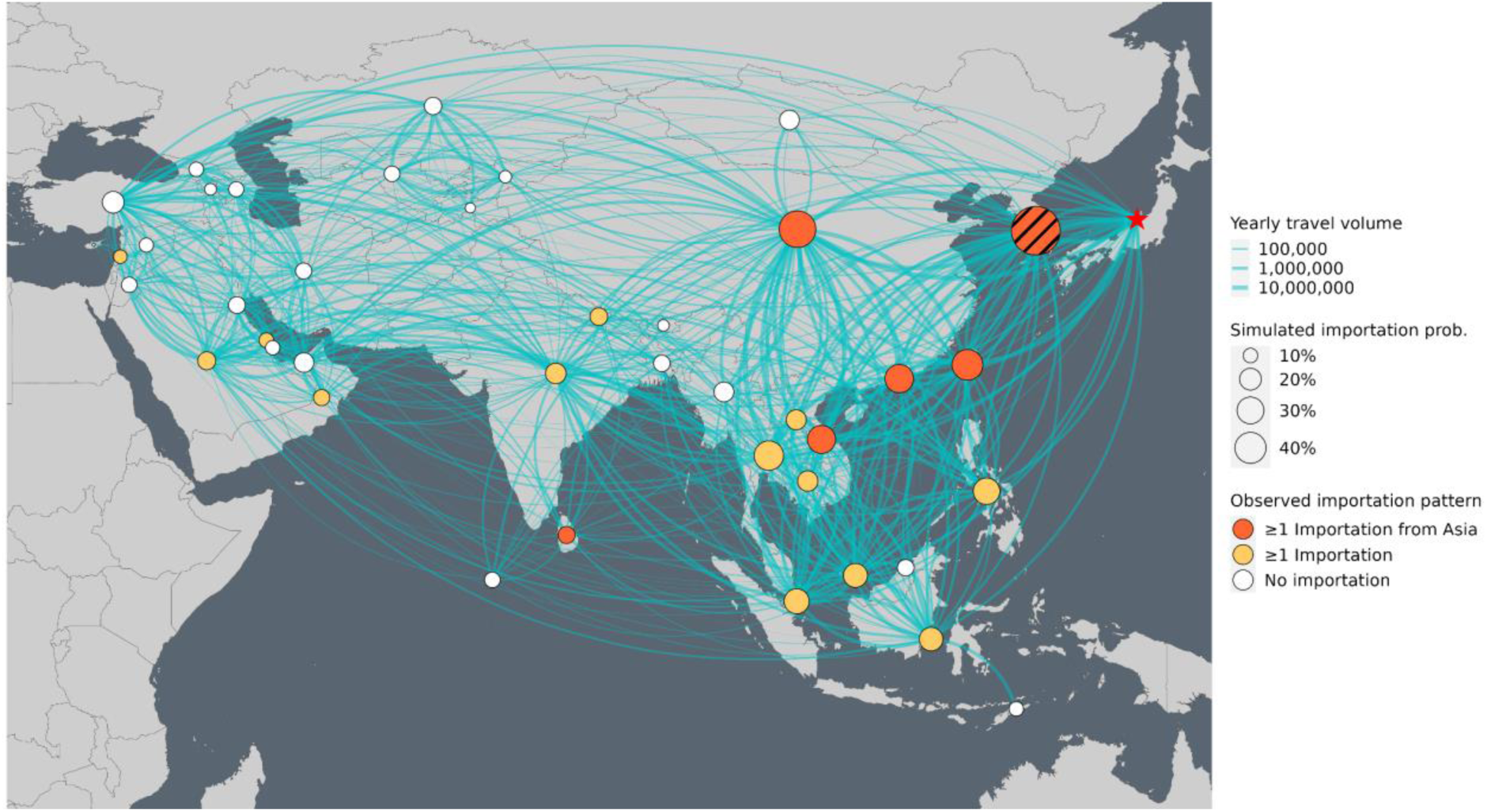
Simulated mpox importation probabilities in Asia. The node size represents the simulated importation probability for each country under the condition of having ≥10 mpox cases in South Korea (hatched node). The widths of links between nodes indicate the average yearly travel volume between countries. The countries with ≥1 known importation from any Asian countries and with ≥1 known importation (either unknown to be from Asia or known to be from outside Asia) are shown in red and yellow, respectively. A red star represents the assumed initial source of spread from Japan. Links for travel volume of less than 10,000 persons per year were omitted from the figure.

### Projected patterns of international spread within Asia

We visualised the international spread patterns of mpox in Asia using an Alluvial diagram (Figure 2). We defined an international spread event as the first occurrence of importation of an mpox case in 2023 between a specific importation-exportation country pair. The boxes and paths in the Alluvial diagram represent the simulated probability of the international spread events and their generational orders (assuming Japan as the initial source of spread, i.e. “Generation 0” in Figure 2) among the simulations. The majority of the 1st-generation importations (i.e. direct importations from Japan) were to Eastern Asian countries, whereas South-eastern Asian countries were predominant in the 2nd generation. The share of Central, Southern and Western Asian countries increased in the 3rd generation onwards, although with relatively small simulated importation probabilities. These simulated patterns generally aligned with reported dates of new mpox introductions in 2023 in Asia (Table S2).

**Figure 2.**
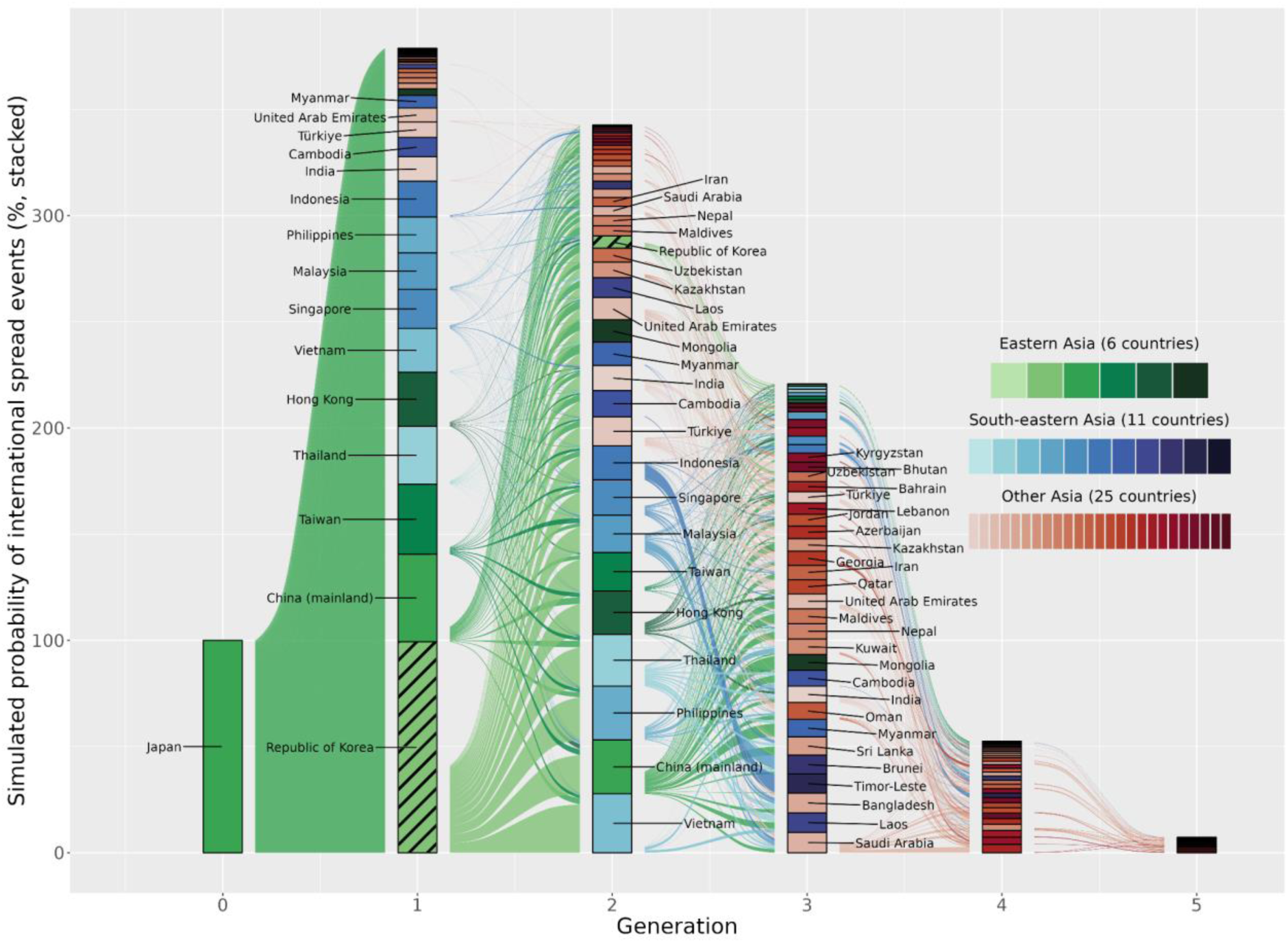
International spread patterns of mpox in Asia. Each column represents the generation of international spread. The height of each box denotes the simulated probability of international spread events for each generation conditional to ≥10 mpox cases in South Korea (hatched boxes). The width of the outflow from the right-hand side of each box (country) denotes the relative frequency of exportation. Green boxes correspond to Eastern Asia, blue boxes to South-eastern Asia and red boxes to Central, Southern and Western Asia.

## Discussion and conclusion

Our multi-country mpox transmission model accounting for heterogeneous sexual contact networks suggested the possible patterns of international spread in Asia. A simulated outbreak was assumed to have originated in Japan, which showed the earliest sustained local outbreak among Asian countries in 2023. Our results highlight that countries at high risk of importation, some of which coincided with the observed importations in 2023, include LMICs mostly located in South-eastern Asia. This is particularly concerning given the large population sizes and limited access to mpox vaccines in these countries. A large outbreak in China in the summer of 2023,^1^ followed by reports of cases in a few neighbouring countries,^10^ was an example of such regional concern. While we note substantial uncertainty in the absolute magnitude of simulated importations due to the assumptions in our model, the relative patterns of importations were suggested to be more robust in our sensitivity analysis (see *Supplementary materials*). We therefore believe that, despite limitations, our simulation would help anticipate the possible future trajectory of mpox international spread in Asia. Indeed, new introductions of mpox cases in 2023 have been reported initially from Eastern Asian countries followed by South-eastern Asian countries, consistent with our simulation results thus far. Whether this further develops into established transmission in South-eastern Asia and then in Central, Southern and Western Asia warrants close attention from the global health community to monitor the risk of global resurgence of mpox.

The 2022 global mpox outbreak has affected many countries, mostly in Europe and the Americas, where it is no longer considered an imminent public health threat^2^ due to likely established immunity through either natural infection or vaccination.^4^ However, there have been multiple events that suggest a potential risk of reemergence, e.g. new cases in areas where the epidemic appeared to have ceased^11^ and reports of breakthrough infections and reinfections.^12^ Even if the transmission potential of mpox has been once depleted due to accumulated immunity in a population, multiple factors including waning immunity, turnover of sexually active individuals or the emergence of immune escaping variants could still drive the reemergence in the long term. Continued circulation of mpox in Asian countries is therefore of concern not only for this region but also for other regions including those that have already been affected in 2022. Global support for control effort in LMICs in Asia, as well as other regions at increased risk, remains essential.

## Author contributions

Conceptualisation: A.E.; Data curation: T.R.A., S-m.J., C.G., H.S., A.T., F.M., A.E.; Formal anlaysis: T.R.A., S-m.J.; Funding acquisition: A.E.; Methodology: T.R.A., S-m.J., H.M., F.M., A.E.; Project administration: A.E.; Software: T.R.A., S-m.J.; Supervision: A.E.; Validation: T.R.A.; Visualisation: T.R.A., S-m.J.; Writing - Original Draft Preparation: T.R.A., S-m.J., A.E.; Writing - Review & Editing: S-m.J., H.M., C.G., H.S., A.T., F.M., A.E.

## Data Availability

All the data used in this study is publicly available, except for the NInJaS study under an ethics-associated restriction on data sharing.

## Material and methods

### Data source

We collected mpox incidence by date of medical attendance in Japan from the Ministry of Health, Labour and Welfare (MHLW) website from 25th July 2022 to 7^th^ July 2023 (https://github.com/akira-endo/mpoxlinelist_JPN). We used the sexual partnership distribution estimated from the self-reported number of sexual partners over 4 weeks of the UK National Survey of Sexual Attitudes and Lifestyles (Natsal) datasets to model the mixing among the MSM population.^1^ We also used the sexual partnership distribution over 1 year estimated from Natsal as part of our sensitivity analysis.^2^ The international travel volume in 2019 was obtained from the World Tourism Organization (UNWTO).^3^

Asian countries included in the present study were based on the United Nations’ definition^4^ but excluded North Korea, Afghanistan, Pakistan, Iraq, Turkmenistan and Yemen due to the missing international travel volume data.

To qualitatively validate our international projections of mpox, we collected the dates of the first mpox report in 2023 in Asian countries from various sources. We also collected the cumulative number of mpox infections from the Our World in Data website^5^ and the weekly incidence of mpox in Japan until the 2023 year-end from the National Institute of Infectious Diseases, Japan.^6^

In addition, we used the National Inventory of Japanese Sexual Behavior (NInJaS) data^7^ to assess the plausibility of our assumption that the sexual partnership distribution in Asia follows the UK Natsal dataset (see Figure S11). The Regional Ethics Committee at the University of Tokyo, Japan approved the study (ref: 2019305NI-(2)).

All the data used in this study is publicly available, except for the NInJaS study under an ethics-associated restriction on data sharing (see the original study^7^ for details).

### Model of international spread of mpox

We developed a stochastic meta-population SEIR model accounting for heterogeneous MSM sexual networks. We focused solely on the MSM populations in our model given the demonstrated predominance of this population group both empirically and theoretically^2,8,9^ in the global mpox outbreak. To reflect the high heterogeneity in empirical data on sexual behaviours,^2^ the MSM population in each country was subdivided into 50 groups of different sexual activity levels (i.e. rates of sexual encounters). Each group was indexed by *k*=1,2,…,50 and is matched with one of the evenly spaced bins on a logarithmic scale between 1 and 10,000 partners per year. We allocated individuals to each group proportionally to the sexual partner distributions estimated from the reported 4-week (or 1-year for the sensitivity analysis) sexual partners in the Natsal data,^1,2^ and the rates of sexual encounters per day for each group, *r*_k_, was calculated as the mean given the probability density within each bin, scaled to the daily unit (Figure S1).

We described the transitions between compartments as a Markov process in daily time steps. We let the daily transitions follow binomial processes:

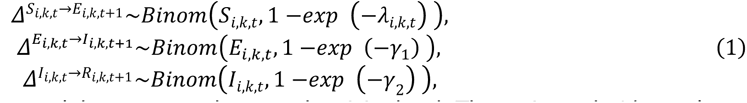

where *i* represents the country and *k* represents the sexual activity level. The reciprocals 1/*γ*_1_ and 1/*γ*_2_ correspond to the assumed latent period (3 days^10^) and infectious period (10 days^11^) of mpox, respectively. The force of infection for those with sexual activity level *k* in country *i* on day *t*, *λ*_i,k,t_, was given as the product of the transmission coefficient *β* and the rate of sexual encounters with infected individuals per day. We modelled the risk of experiencing infectious sexual contacts for individuals residing in country *i* considering three different transmission pathways: local transmission in country *i*, transmission from local individuals in country *j* whilst outbound travels, and transmission from inbound travellers visiting from country *j*.

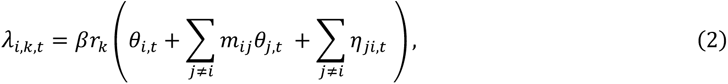

where the first term (βr_k_*Θ*_i,t_) represents the daily rate of local individuals with sexual activity level *k* being infected by a local infectious sexual partner in country *i* on day *t*, the second term represents the daily rate of travellers from country *i* getting infected outside of country *i* on day *t*, and the third term represents the daily rate of local individuals being infected by travellers visiting from outside of country *i* on day *t*. Here, *Θ*_i,t_ represents the risk of contacting infectious sexual partners in country *i* and *η*_ji,t_ represents the risk of contacting infectious sexual partners in country *i* who are inbound travellers from country *j*. Assuming proportionate mixing, these risks were modelled as the proportion of infectious sexual contacts on day *t* (*C*_inf,i,t_) among the overall sexual contacts (*C*_all,i,t_) over the infectious period 1/*γ*_2_.

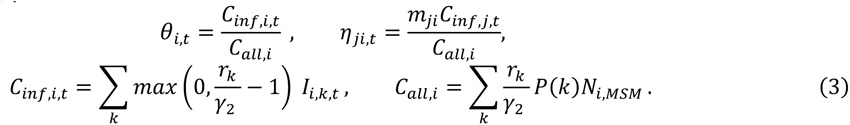

*N*_i,MSM_ is the MSM population size in country *i*. When constructing *C*_inf,i,t_, we subtracted one from the number of sexual partners that an infectious person is expected to have over the infectious period of 1/*γ*_2_. This is to reflect the concept of the excess degree,^12^ which assumes that the number of partners one can transmit to is the total number of partners except one who infected the person. For interpretability, we transformed *β* into the secondary attack risk (SAR), i.e. transmission risk per sexual partner: SAR = 1-exp(-β).

International mixing matrix *m*_ij_ represents the cross-sectional proportion of individuals from country *i* travelling in country *j* and is the product of two factors: the daily number of travellers per capita and the average duration of international travels, *D*_T_, which we assumed to be 7 days.

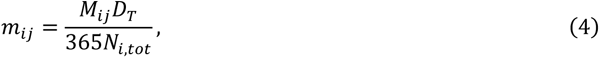

where *M*_ij_ is the annual travel volume from country *i* to country *j*, while *N*_i,tot_ is the total population size in country *i*. It is noted that we used the international travel volume in 2019, which was not affected by the COVID-19 restriction.

During the simulation of the stochastic meta-population model described in Equation (1), we also sampled the daily number of importations between countries in a post-hoc manner. Specifically, we first sampled the total number of importations on day *t+1* from any country, *n*_i,imp_(t+1), among new infections 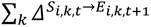.

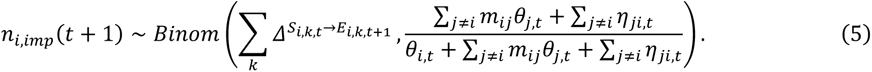

Then, if *n*_i,imp_(t+1) is nonzero, the number of importations stratified by the source country *j*, *n*_i,imp,j_(t+1) was determined by multinomial sampling:

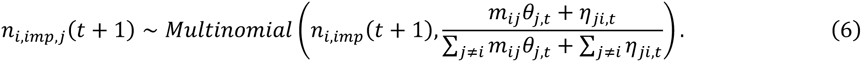

### Model parameter specifications

We summarise the parameter values used in the present study in Table S1.

### Model fitting using alive particle filter

We calibrated our stochastic SEIR model for Japan to the observed weekly incidence from 16^th^ January to 26^th^ June 2023 by medical attendance date to estimate the transmission coefficient *β*. Where the medical attendance date was missing, we imputed it from the reporting date, using the mean empirical delay from medical attendance to reporting within our data, which we estimated to be 9 days. We assumed that the effect of intervention or behaviour change was negligible during this period. We used a Bayesian data assimilation approach via the alive particle filter (APF), one of the approximate Bayesian computation (ABC) algorithms.^14,15^ This approach allowed us to obtain posterior samples of *β* alongside the corresponding trajectories of mpox incidence by sexual activity levels *k* in Japan.

Following McKinley et al.,^14^ we implemented the ABC pseudo-marginal Metropolis-Hastings (ABC-PMMH) algorithm in which the likelihood was approximated by the APF. For efficiency, we employed two types of early rejection, which are described in section 4 in McKinley et al.^14^ First, a proposal was rejected if the total number of simulations in the APF exceeded the pre-specified number, which we set as 5,000 multiplied by the number of particles (300 in our case). Second, during each iteration of the ABC-PMMH, the simulation may be terminated part-way to reject the proposal based on the interim likelihood where possible.

For the implementation of ABC, we employed the following tolerance *ε*_t_ based on the observed weekly incidence (*y*_t_). Let the function *h*_t_ be

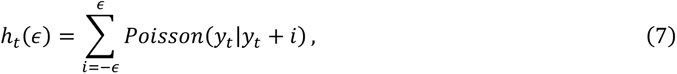

and we defined *ε*_t_ as the smallest *ε* that satisfies *h*_t_(*ε*) ≥*p*_0_, where *p*_0_ is a pre-specified threshold (set at 0.5 in our study). The absolute difference between the simulated weekly incidence (*x*_t_) and *y*_t_ should be less than *ε*_t_ at every time point *t* for acceptance.

We ran the model simulation initiating with one mpox case in Japan (corresponding to the first reported case with a medical attendance date of 16^th^ January 2023) within the APF. The sexual activity level *k* for this initial case, for each run, was sampled from the sexual degree distribution *P(k)* weighted by the degree, which we assumed to be proportional to the risk of infection, as shown in Figure S1B&C:

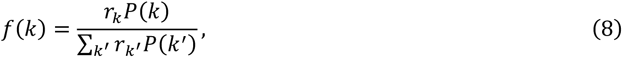

This distribution *f(k)* corresponds to *f*_*x*_*_0_* (· |θ) in Algorithm 1 in McKinley et al.^14^

We used a weakly-informative prior for *β* (a half-Cauchy distribution with a location parameter of 0 and a scale parameter of 2.5). We obtained 10,000 samples from a single chain of ABC-PMMH after discarding the first 2,000 samples as burn-in, which were then thinned to 1,000 samples to reduce auto-correlation. We ensured that the effective sample size was at least 200 and the R-hat value was less than 1.05 for all chains.^16^ We ran the APF with the obtained posteriors of *β* to record the associated trajectories of mpox infections.

### Simulating international spread

Our ABC-PMMH model fitting to incidence data in Japan yielded 1,000 samples of transmission coefficient *β* and associated trajectories of mpox infections (i.e. the number of individuals in the SEIR compartments stratified by sexual activity level *k*) in Japan until 26 June 2023. For consistency with the observed international spread in Asia, we further conditioned these samples of *β* and trajectories on the established importation in the Republic of Korea (South Korea) by the end of the fitting period (26 June 2023). We first simulated international spread in Asian countries using our metapopulation SEIR model given the previously obtained (unconditional) samples of *β* and infection trajectories. That is, we ran the model with prespecified *β* and incidence in Japan to obtain 50,000 simulations (50 runs per unconditional sample) of mpox international trajectories over three years (which were long enough to observe the entire course of spread in considered countries). We then limited these simulations to only those observing ≥10 mpox cases in South Korea by the end of the fitting period. We recorded the first importation between specific country pairs in each simulation run to visualise the simulated probabilities of importations and their generational orders.

All the analysis was performed in Julia v1.8.3. Visualisation was done in Julia v1.8.3 and R v4.2.2. We used the R package {ggalluvial} v0.12.5 to draw the Alluvial diagram. The data and codes are deposited at https://github.com/toshiakiasakura/projection_of_mpox_in_Asia.

## Supplementary results

### Epidemic situation in Japan

In the earlier phase of the 2022 global mpox outbreak when Western countries were particularly affected, only sporadic cases were observed in most Asian countries, including Japan. From 16^th^ January 2023 onward, sustained local transmission was reported in Japan. Among those cases in 2023, three were known to have reported international travel history (Figure S2). Two visited Asia and one visited North and Central America.

### Projected mpox incidence in Japan

We projected the international spread of mpox, and we visualised the projected mpox incidence with and without the condition that ≥10 cases were observed in South Korea during the fitting period and compared it with the mpox incidence in Japan in 2023^6^ (Figure S3).

### First mpox reporting dates and cumulative cases in 2023 in Asian countries

We collected the first date of the mpox report in 2023 in each Asian country as of 8 April 2024 and the cumulative incidence of mpox as of 3 March 2024 from the Our World in Data (OWID) websites^5^ (Table S2). If we did not find a citable source on the first case report, we used the first reporting date as recorded in OWID.

Countries without any reported cases as of 31 December 2023 are not included in Table S2: 1 out of 6 in Eastern Asian countries (Mongolia); 3 out of 11 South-eastern Asian countries (Brunei, Timor-Leste, and Myanmar); 4 out of 7 Southern Asian countries (Bangladesh, Bhutan, Maldives, and Nepal); all the 4 Central Asian countries (Kazakhstan, Kyrgyzstan, Turkmenistan and Uzbekistan); 4 out of 14 Western Asian countries (Syria, Kuwait, Azerbaijan and Armenia).

According to OWID data, the mpox outbreak size in 2023 is 3,332 and the median of projected final outbreak size in Asian countries is 231 (95% credible interval: 183, 971057). The number of countries with any reported cases between 1st January and 31st December 2023 is 18 excluding Japan, and the median of the projected countries experiencing importation is 4 (95% credible interval: 2, 41).

### Sensitivity analysis

As part of the sensitivity analysis, we estimated the SARs using the same approach as the main analysis but with varying assumptions for the sexual partner distribution and infectious period for the sensitivity analysis (Table S3). Specifically, instead of the reported number of 4-week sexual partners from the Natsal data we used for the main analysis, we used the number of 1-year sexual partners as the reference for the partnership distribution over the infectious period of mpox. We used the parameter values for the Weibull distribution, *σ* = 0.10 and *κ* = 0.77, estimated in the previous study (Table S1).^2^ We also varied the assumed infectious period (7 days or 14 days, instead of 10 days in the main analysis). Figures S4–S7 show the trace plot and posterior distribution for each scenario.

Using the same approach as the main analysis, we projected the international spread of mpox for the additional three scenarios considered in our sensitivity analysis. We show the simulated importation probabilities, along with the distribution of final outbreak size (i.e. total cumulative case counts at the end of the simulation) in Asia and the number of countries with at least one mpox case among the simulations (Figure S8). We also compared the simulated and observed first importation dates (observed reported dates only include those known as of the end of 2023) (Figure S9). Finally, we varied the conditionals on the international spread we used in model fitting. In the main analysis, we conditioned the posterior distribution such that there were at least 10 cases in South Korea in the simulations by the end of the fitting period. Instead, we applied three alternative conditions: (i) unconditional (model fitted solely based on incidence in Japan); (ii) conditional to ≥10 cases in Taiwan; and (iii) conditional to ≥10 cases in mainland China (Figure S10).

### Comparison of the sexual partnership distribution between the UK and Japan

To assess the plausibility of the assumption that the sexual partnership distribution in all considered countries follows the empirical data from the UK (Natsal), we compared the Natsal’s 1-year distribution with that from the National Inventory of Japanese Sexual Behavior (NInJaS) collected in Japan.^7^

The sexual partnership distribution among MSM used in part of our sensitivity analysis, based on the UK Natsal data,^2^ was derived from the reported number of sexual partners among men aged 18-44 who reported at least one same-sex sexual partner over a year. The exactly comparable data was not available in the NInJaS data, which collected the number of sexual partners in the last 12 months among men aged 20-39 reporting having sex with men in their lifetime but without exactly specifying the gender of partners. That is, the participants in NInJaS reported the number of overall sexual partners over 12 months and whether these partners were, as a whole, (i) all opposite-sex, (ii) mostly opposite-sex, (iii) half-and-half, (iv) mostly same-sex or (v) all same-sex. We attempted to infer the distribution of the number of same-sex partners among MSM in the NInJaS data by assigning weights to the reported number of partners according to the reported breakdown of gender (i)–(v); i.e. when a participant reported *n*_p_ sexual partners in the previous year, we assumed that the following fraction *p*_*M*_ among these partners was male: (i): *p*_*M*_ = *0*; (ii): *p*_*M*_ = *ψ*; (iii): *p*_*M*_ = *0*.*5*; with a response (iv): *p*_*M*_ = *1* − *ψ*; (v): *p*_*M*_ = *1*. We then used *n*_*MSM*_ = *p*_*M*_*n*_*p*_ as the number of male sexual partners among MSM. We generally allowed *n*_*MSM*_ to be non-integer for convenience. However, for comparability with the Natsal data, from which we only included individuals with one or more male sexual partners, we substituted *n*_*MSM*_ ’s that are smaller than 1 with 1’s but with a sample weight of *n*_*MSM*_ (e.g. a sample of *n*_*MSM*_ = *0*.*1* was considered as a sample with one partner and a sample weight of 0.1).

We compared the sexual partner distributions from the Natsal and the NInJaS with *ψ*=0.05, 0.2, 0.4 (Figure S11). Overall, the distributions from NInJaS data showed a similar trend to the Natsal data.

---

**Figure S1.**
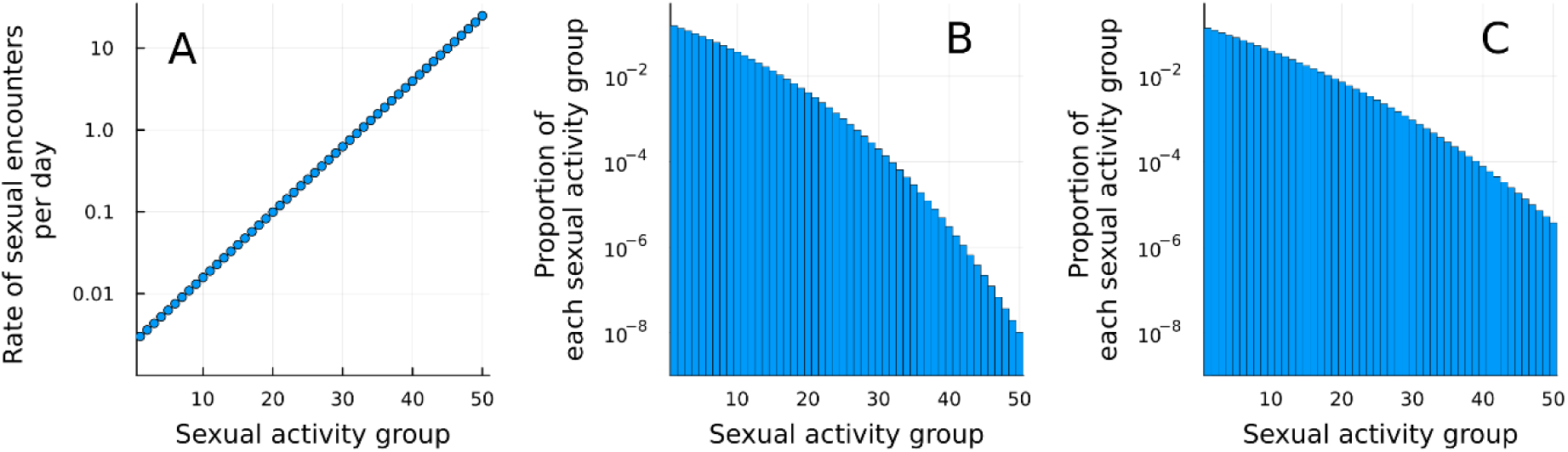
Sexual activity groups for the mpox transmission model over MSM networks. (A) Rate of sexual encounters for each sexual activity group. (B&C) Distribution of sexual activity groups based on the number of (B) 4-week and (C) 1-year sexual partners reported in the Natsal data.

**Figure S2.**
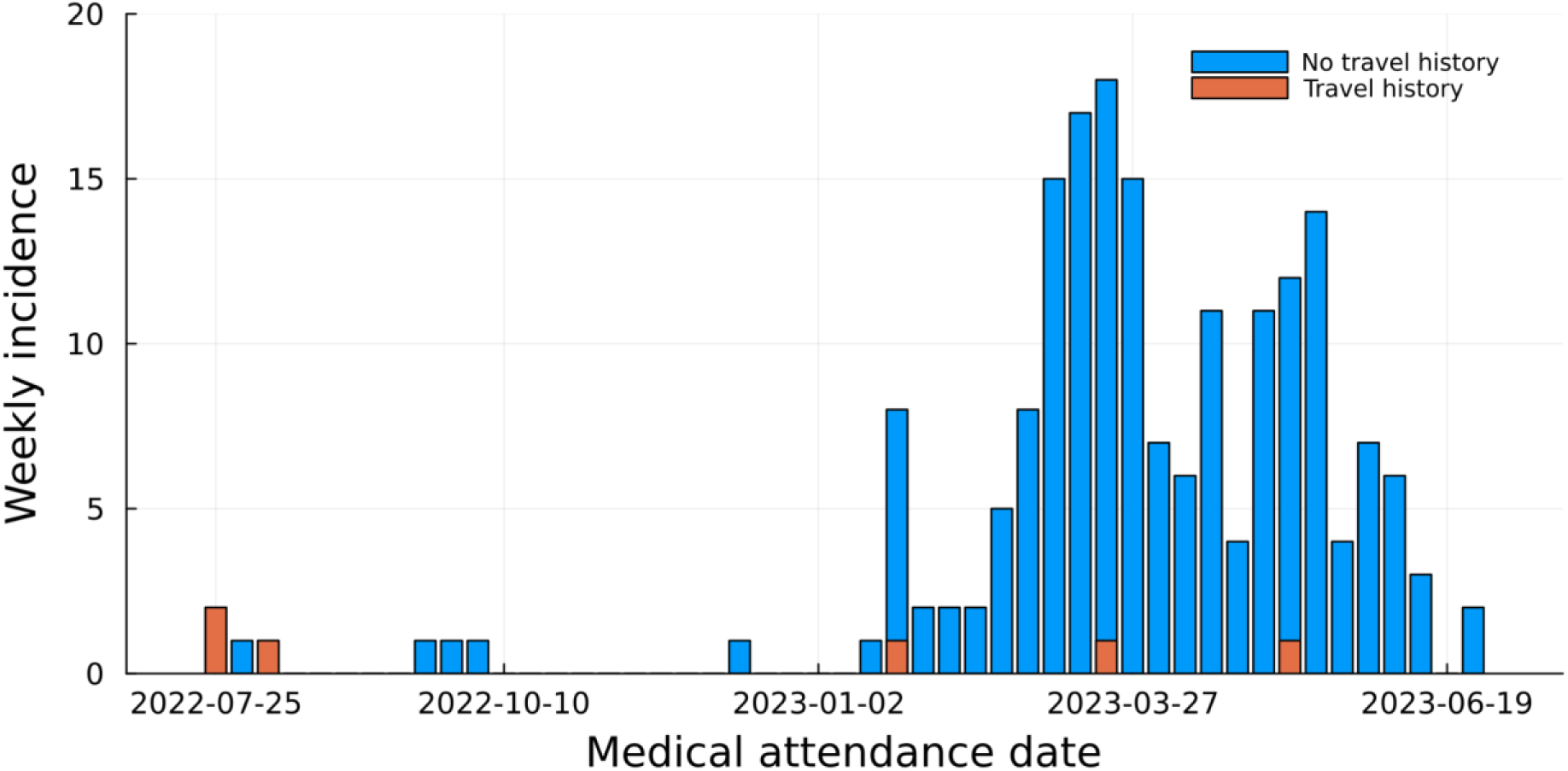
The epidemic curve in Japan, 25 July 2022—26^th^ June 2023.

**Figure S3.**
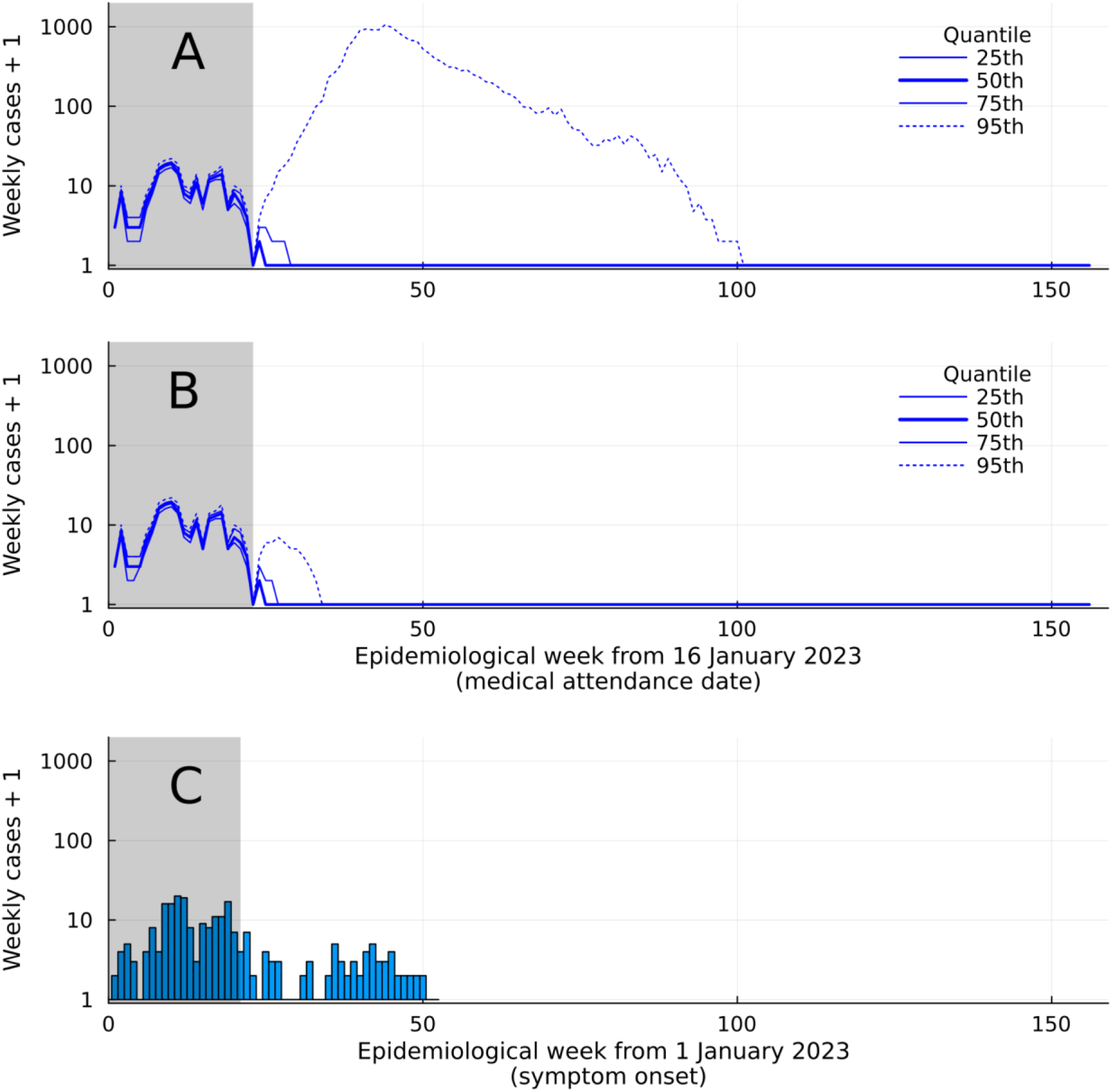
Projected mpox incidence in Japan from posterior samples. (A) The simulated trajectories of mpox incidence from the posterior samples conditioned on ≥10 cases in South Korea The shaded area represents the fitting period from 16^th^ January to 26^th^ June 2023. (B) The simulated trajectories of mpox incidence from unconditional posterior samples (C) Observed mpox incidence in Japan in 2023 (until the 2023 year-end, i.e. week 52).

**Figure S4.**
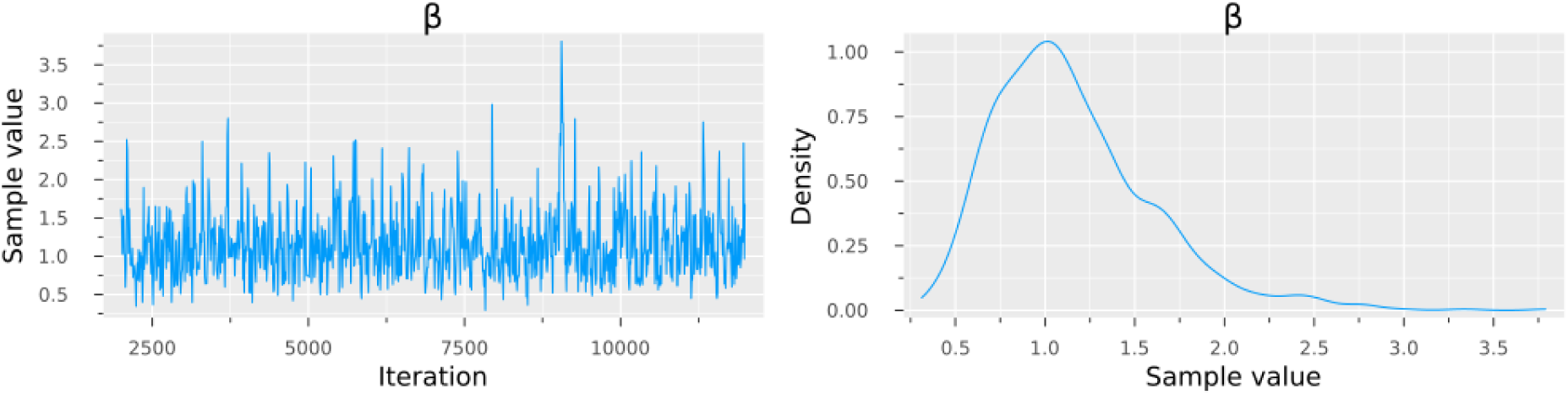
ABC-PMMH trace plot and the posterior distribution of *β* for scenario 1 (main analysis). We assumed the sexual partner distribution based on the reported 4-week number and infectious period of 10 days (main analysis).

**Figure S5.**
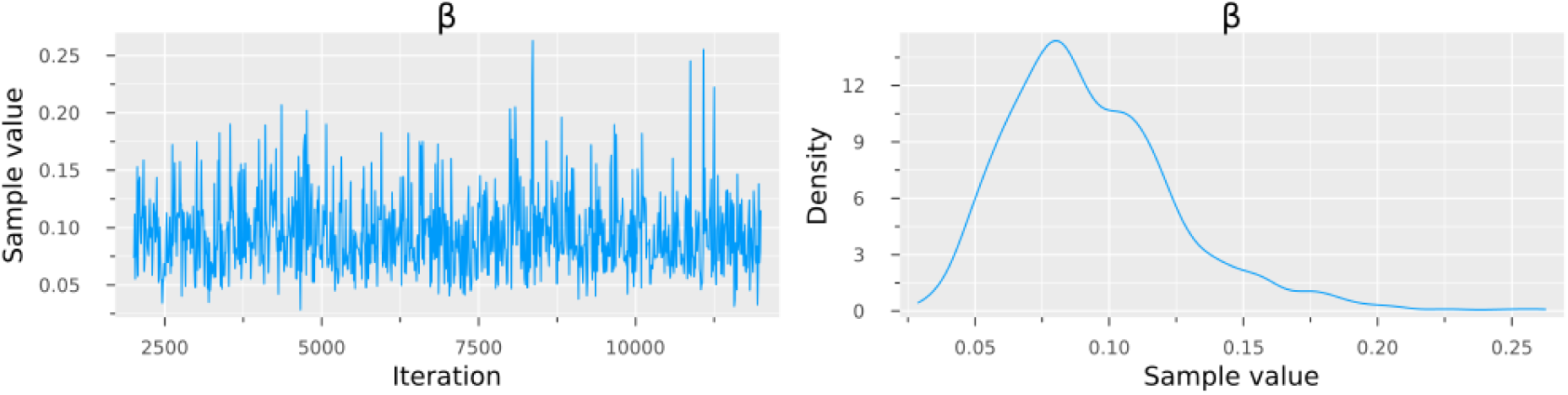
ABC-PMMH trace plot and the posterior distribution of *β* for scenario 2. We assumed the sexual partner distribution based on the reported 1-year number and infectious period of 10 days.

**Figure S6.**
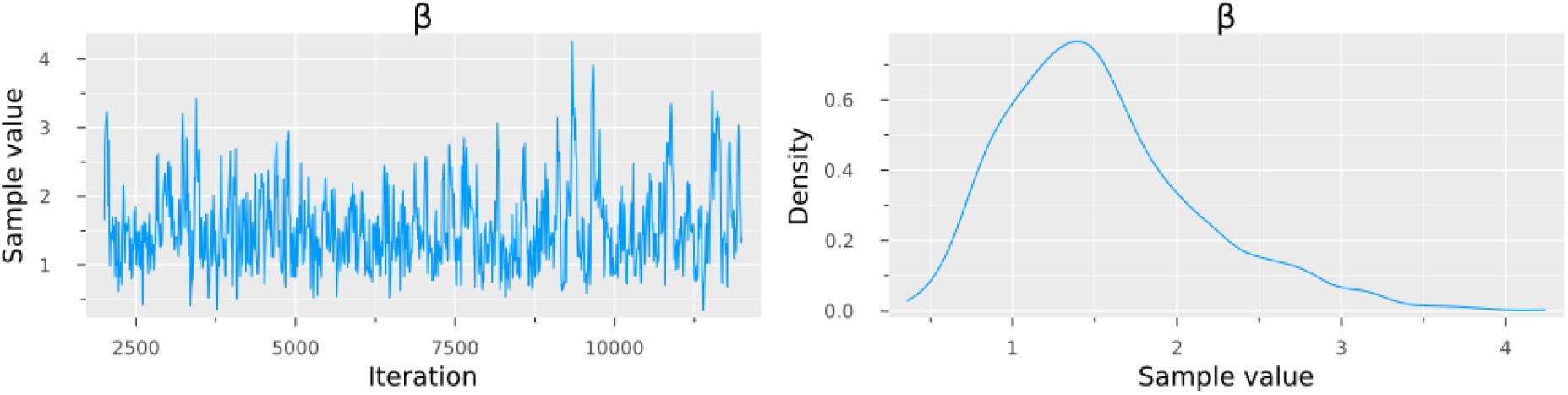
ABC-PMMH trace plot and the posterior distribution of *β* for scenario 3. We assumed the sexual partner distribution based on the reported 4-week number and infectious period of 7 days.

**Figure S7.**
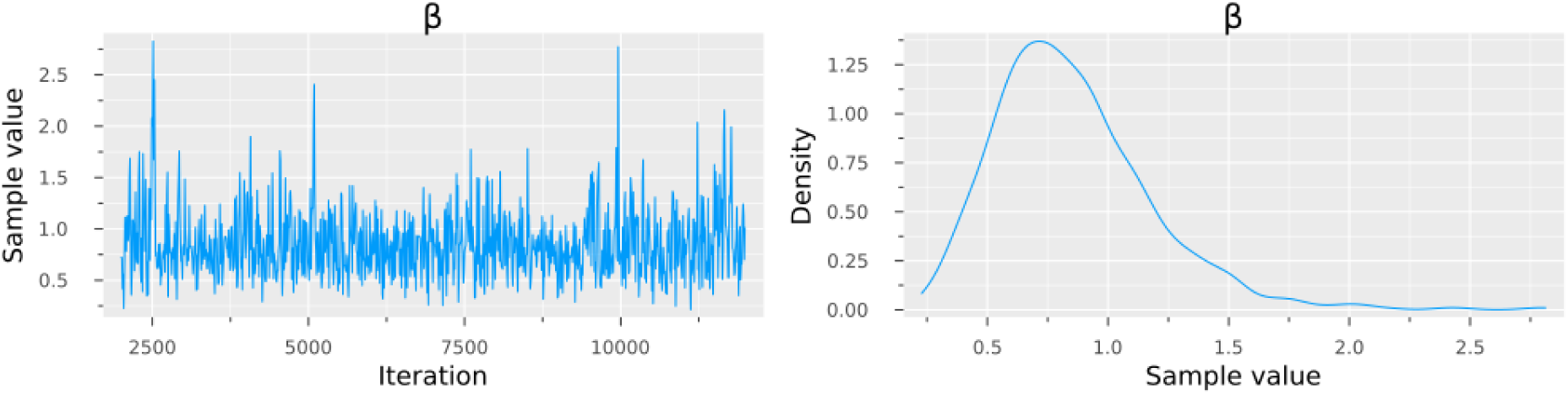
ABC-PMMH trace plot and the posterior distribution of *β* for scenario 4. We assumed the sexual partner distribution based on the reported 4-week number and infectious period of 14 days.

**Figure S8.**
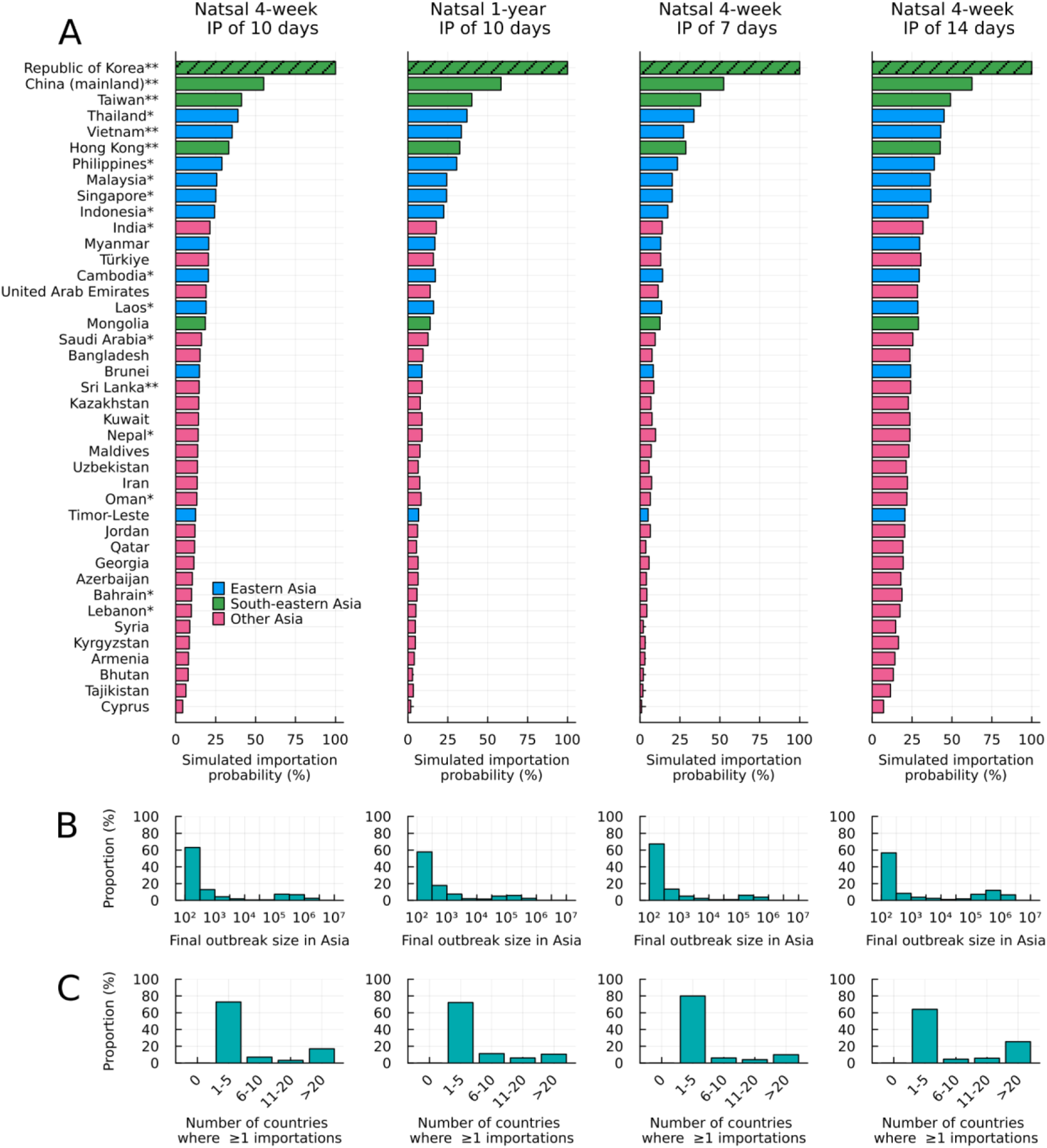
Projected outcomes from the international spread model for four scenarios. (A) The simulated importation probabilities conditioned on ≥10 cases in South Korea (hatched bars). Double asterisks (**) beside country names indicate ≥1 importation known to be from other Asian countries and a single asterisk (*) indicates≥1 importation but not known to be from Asia. (B) Distribution of the final outbreak sizes in Asia among the simulations, defined as the total number of cases over all the Asian countries included until the end of simulations. (C) Distribution of the number of countries with ≥1 importation. IP: infectious period.

**Figure S9.**
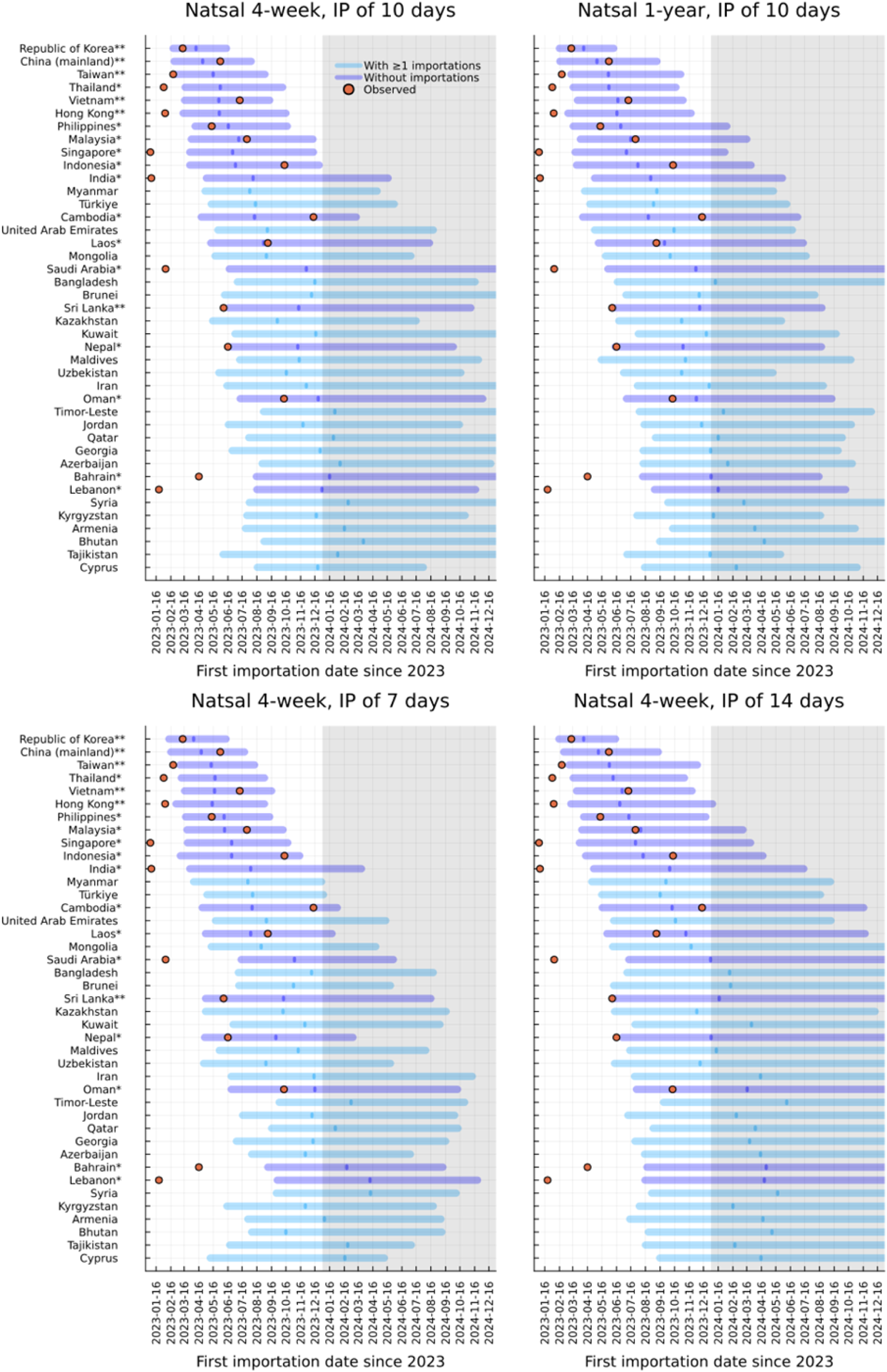
Simulated and observed first importation dates. The simulated (bars) and observed (red circles) first importation dates for mpox in Asia by date of reporting. Observed first importation dates were restricted to those reported by the end of 2023 (indicated by the grey shade). Simulated importation dates were assumed to be lagged 12 days from the dates of infection. The simulations were run on 16 January 2023 onward. The horizontal bars and the vertical lines in their middle represent 90% simulation ranges and the medians for the first importation dates. Different bar colours distinguish between countries with (blue) and without (light blue) ≥1 observed importation in the real world as of 2023 year-end. Shaded areas from 2024 represent periods when the 1^st^ importation dates were not yet collected or unavailable. Double asterisks (**) beside country names indicate ≥1 importation known to be from other Asian countries and a single asterisk (*) indicates≥1 importation but not known to be from Asia. IP: infectious period.

**Figure S10.**
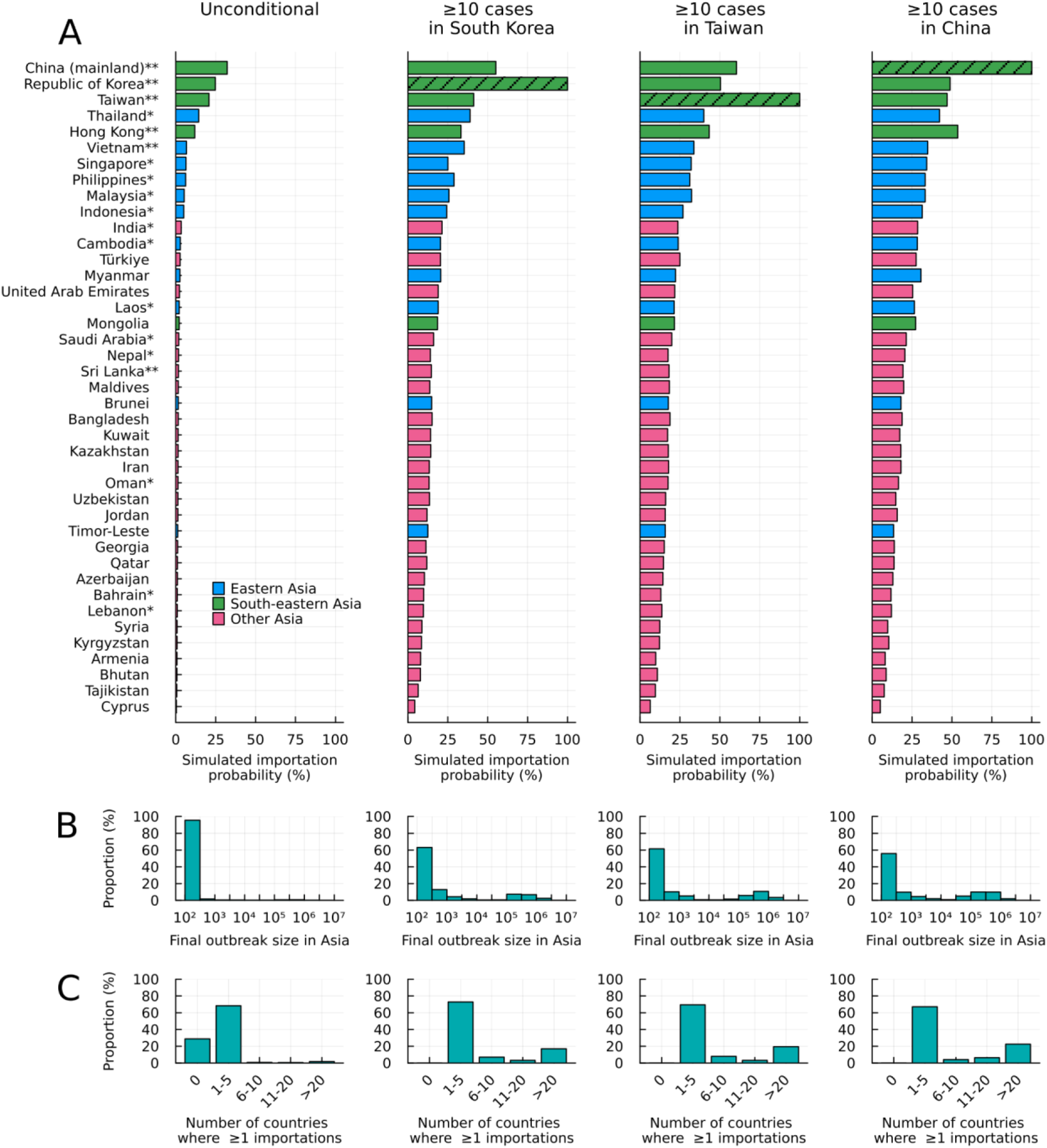
Projected outcomes from the international spread model for different conditions. (A) The simulated importation probabilities for alternative conditionals. Hatched bars represent the country on which the estimation was conditioned (to have ≥10 cases). Double asterisks (**) beside country names indicate ≥1 importation known to be from other Asian countries and a single asterisk (*) indicates≥1 importation but not known to be from Asia. (B) Distribution of the final outbreak sizes in Asia among the simulations, defined as the total number of cases over all the Asian countries included until the end of simulations. (C) Distribution of the number of countries with ≥1 importation. IP: infectious period.

**Figure S11.**
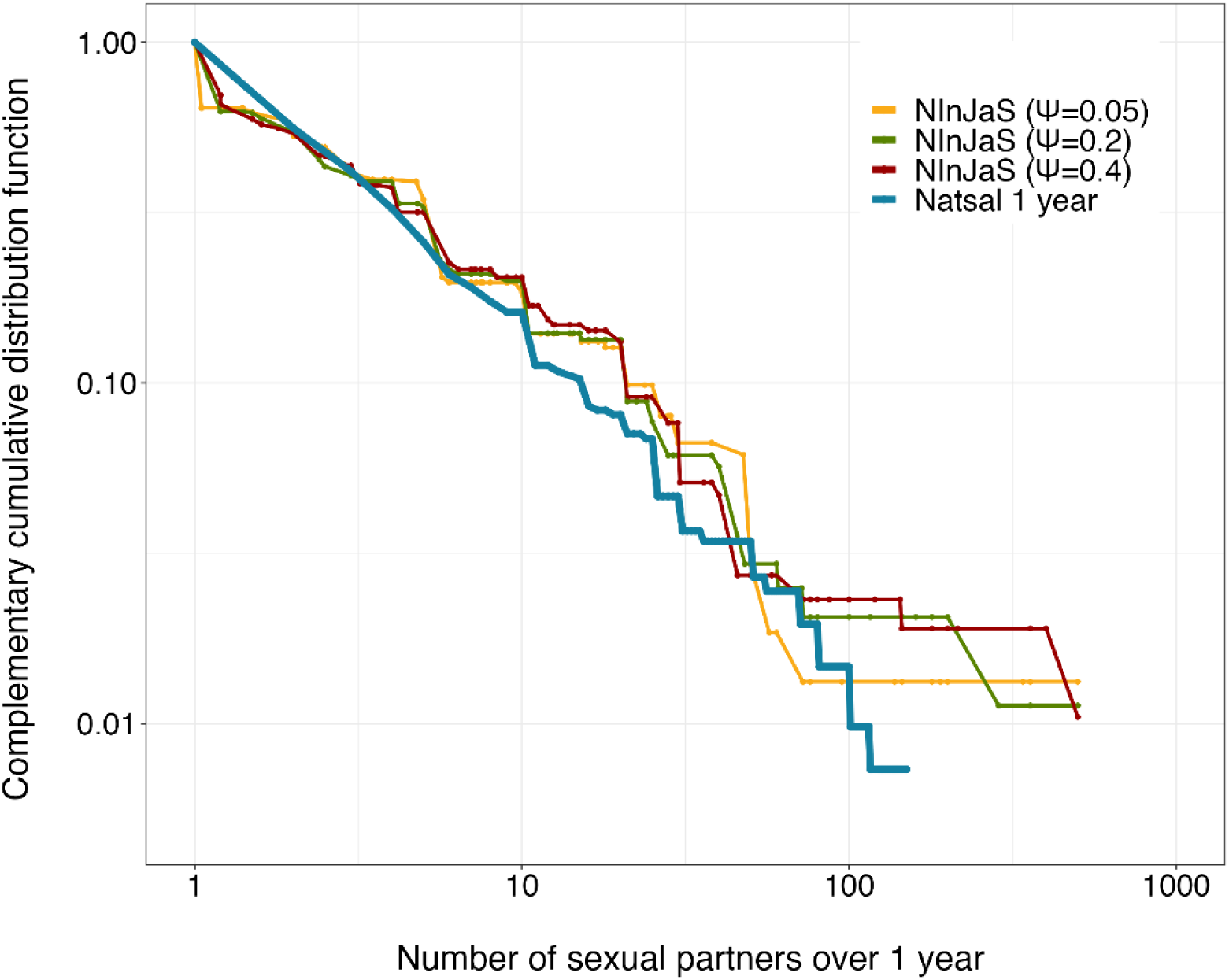
The complementary cumulative distribution function for the 1-year sexual partnership distribution for the Natsal and NInJaS datasets. The parameter *ψ* represents the assumed proportion of males among the reported sexual partners when the respondents indicated their partners were mostly opposite-sex / same-sex.

**Table S1.**
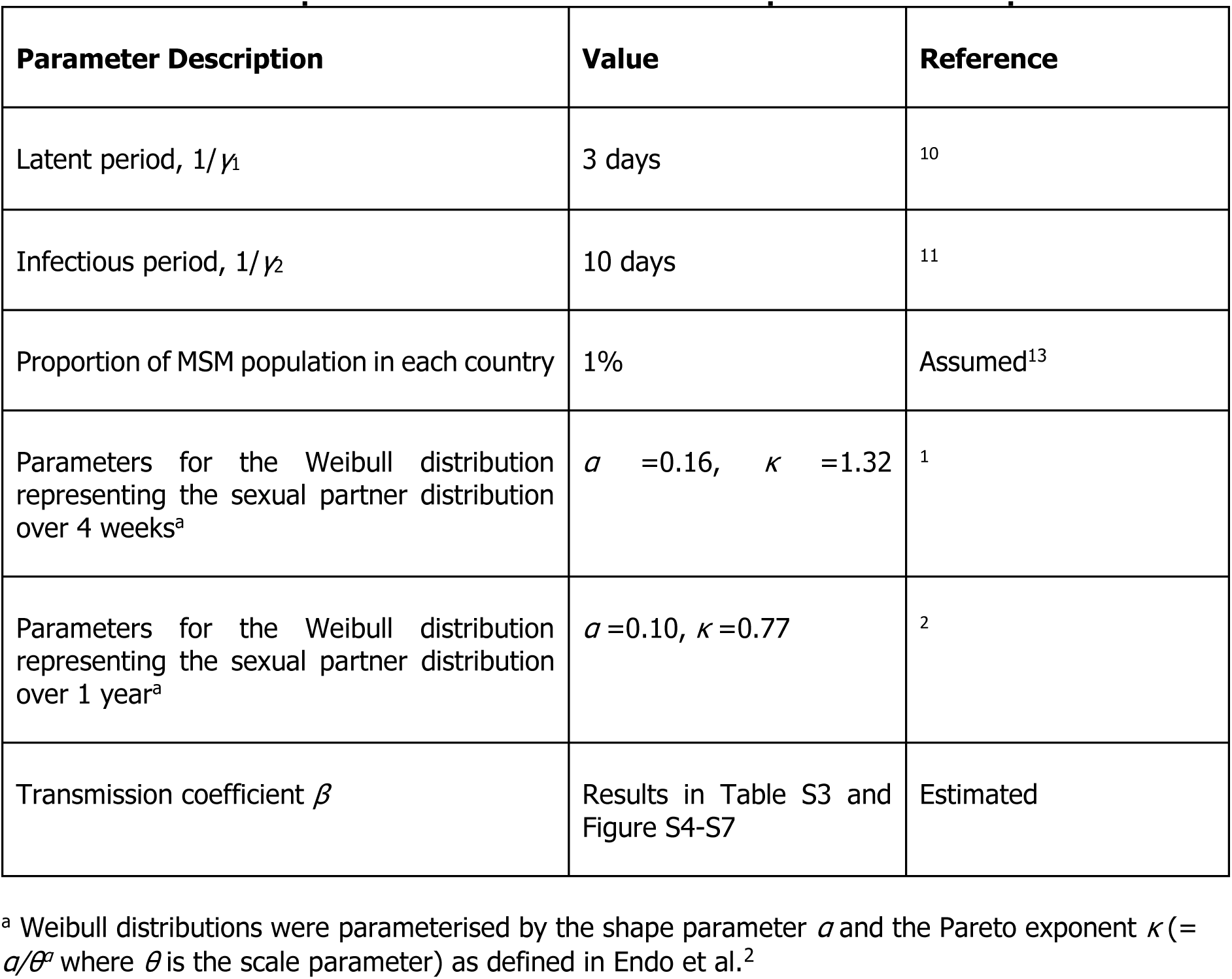
Parameter specification for the international spread model of mpox.

**Table S2.** Cumulative incidence for 2022 and 2023, and the first reporting date of mpox in Asian countries (data as of 8 April 2024).

| UN Region | Country | Cumulative incidence in 2022 | Cumulative incidence in 2023 | First reporting date in 2023 | Source for the first report in 2023 |
| --- | --- | --- | --- | --- | --- |
| Eastern Asia | Japan | 8 | 224 | 2023-01-19* | 17,18 |
| Eastern Asia | Hong Kong | 1 | 53 | 2023-02-04* | 19 |
| Eastern Asia | Taiwan | 4<sup>a</sup> | 355<sup>a</sup> | 2023-02-15* | 20–22 |
| Eastern Asia | Republic of Korea | 4<sup>a</sup> | 151<sup>a</sup> | 2023-03-13* | 23,24 |
| Eastern Asia | China (mainland) | 1<sup>a</sup> | 1611<sup>a</sup> | 2023-05-31* | 25 |
| South-eastern Asia | Singapore | 19 | 31 | 2023-01-01 ~ 2023-01-07<sup>b</sup> | 26 |
| South-eastern Asia | Thailand | 12 | 673 | 2023-02-01 | OWID<sup>c</sup> |
| South-eastern Asia | Philippines | 4 | 5 | 2023-05-13 | OWID<sup>c</sup> |
| South-eastern Asia | Vietnam | 2 | 119 | 2023-07-11* | <sup>27</sup> OWID<sup>c</sup> |
| South-eastern Asia | Malaysia | 0 | 9 | 2023-07-26 | <sup>28</sup> |
| South-eastern Asia | Laos | 0 | 1 | 2023-09-08 | <sup>29</sup> |
| South-eastern Asia | Indonesia | 1 | 72 | 2023-10-13 | <sup>30</sup> |
| South-eastern Asia | Cambodia | 0 | 1 | 2023-12-13 | <sup>31</sup> |
| Southern Asia | India | 20 | 7 | 2023-01-06 | <sup>32</sup> |
| Southern Asia | Sri Lanka | 2 | 2 | 2023-06-07* | <sup>33</sup> |
| Southern Asia | Nepal | 0 | 1 | 2023-06-16 | <sup>34</sup> |
| Southern Asia | Iran | 1 | 0 |  |  |
| Western Asia | Lebanon | 24 | 3 | 2023-01-22 | OWID<sup>c</sup> |
| Western Asia | Bahrain | 1 | 1 | 2023-04-16 | OWID<sup>c</sup> |
| Western Asia | Qatar | 5 | 0 |  |  |
| Western Asia | United Arab Emirates | 16 | 0 |  |  |
| Western Asia | Jordan | 1 | 0 |  |  |
| Western Asia | Georgia | 2 | 0 |  |  |
| Western Asia | Cyprus | 5 | 0 |  |  |
| Western Asia | Oman | 0 | 3 | 2023-10-12 | OWID<sup>c</sup> |
| Western Asia | Saudi Arabia | 8 | 0 | 2023-02-25 | <sup>35</sup> |
| Western Asia | Türkiye | 12 | 0 |  |  |

\* Countries with at least one known importation from other Asian countries.
<sup>a</sup> We collected data for Taiwan and Hong Kong from the following websites.<sup>19,20</sup> For mainland China, we subtracted the cumulative incidence in overall China from those in Taiwan and Hong Kong.
<sup>b</sup> Only the weekly incidence was available and we could not identify the exact reporting date.
<sup>c</sup> We used the first reporting date in 2023 in OWID when we did not find a citable source on the first case report.

**Table S3.** Estimated SARs for each sexual partner distribution and infectious period.

| Scenario | Simulation pattern |  | Unconditional estimates from the ABC-PMMH algorithm |  | Conditional estimates |  |  |
| --- | --- | --- | --- | --- | --- | --- | --- |
| | Sexual partner distribution | Infectious period | $\beta$ (95% CrI.)<sup>a</sup> | SAR, % (95 CrI.) | Valid samples (/ 50,000) | $\beta$ (95% CrI.) | SAR, % (95 CrI.) |
| 1 (main analysis) | Natsal 4-week | 10 days | 1.08 (0.52, 2.31) | 66.0 (40.5, 90.1) | 626 | 1.27 (0.64, 2.65) | 72.0 (47.2, 93.0) |
| 2 | Natsal 1-year | 10 days | 0.09 (0.05, 0.18) | 8.4 (4.6, 16.1) | 856 | 0.10 (0.05, 0.22) | 9.7 (5.1, 19.9) |
| 3 | Natsal 4-week | 7 days | 1.45 (0.66, 3.04) | 76.6 (48.5, 95.2) | 494 | 1.66 (0.81, 3.33) | 80.9 (55.7, 96.4) |
| 4 | Natsal 4-week | 14 days | 0.80 (0.36, 1.56) | 55.1 (30.3, 79.0) | 772 | 0.93 (0.43, 2.01) | 60.5 (34.7, 86.6) |

<sup>a</sup> CrI: credible interval.

